# Assessing Neurological Outcomes at One-Year Corrected Age in Preterms ≤34 Weeks Gestation: Research Protocol for a Prospective Cohort Study

**DOI:** 10.1101/2025.10.01.25337111

**Authors:** Florence Murila, Fred Were, Jalemba Aluvaala, Moses Obimbo

## Abstract

Preterm infants are at high risk of neurodevelopmental disorders, growth faltering, recurrent infections, re-hospitalization, and visual and auditory impairments. Despite the rising burden of preterm births, morbidities and mortalities in sub-Saharan Africa, data on neurodevelopmental and growth outcomes and survival during the first year of life remain scarce. This protocol aims to develop a comprehensive follow-up of care and early screening guidelines for this vulnerable population. The study will determine the prevalence of neurological disabilities at one year of corrected age (CA) among preterm infants (≤34 weeks) discharged from two public hospitals in Nairobi, Kenya. It will identify maternal, neonatal, and clinical factors associated with neurological outcomes, characterize growth patterns and their relationship with neurodevelopmental outcomes and evaluate mortality risk at one year of CA.

This prospective cohort will recruit 420 eligible preterm infants and follow them to one-year CA or death. Follow up assessments at 40 weeks’ post-menstrual age (PMA) or 2 weeks post discharge, at 3, 6, 9, and 12 months of CA will document growth, neurodevelopment using the Ages and Stages Questionnaire (ASQ), and sensory outcomes (hearing and visual). Near Infrared Spectroscopy (NIRS) will be performed at one year of CA. Longitudinal analyses using mixed-effects models and generalized estimating equations will examine growth and neurodevelopmental trajectories, while Cox proportional hazards models will assess mortality risk. The study is approved by the Kenyatta National Hospital-University of Nairobi Ethics (KNH-UoN) Committee, Reference Number P466/05/2023.

This is the first cohort in Kenya to integrate neurological, growth, and sensory outcomes with advanced imaging in preterm infants. Findings will guide the development of an evidence-based care bundle for comprehensive follow-up and early intervention, potentially improving survival and long-term outcomes for this high-risk population, especially in low-resource settings.

## Introduction

World Health Organization (WHO) defines a preterm birth as a live birth that occurs before 37 completed weeks of pregnancy. Preterm birth is of global concern. Data obtained in 2010 from 184 countries informs us that 15 million babies are born prematurely every year globally translating to a preterm birth rate of approximately 11% to 18%.^1^ Data obtained from 103 countries in 2020 showed that 13.4 million newborn babies were born preterm, with Asia and sub-Saharan Africa accounting for around 65% preterm births.^2^

For the baby born prematurely, various studies acknowledge that development and catch-up growth is of uttermost importance during the first 6 months after birth.^3,4^ Studies have shown considerable faltering in growth during this period, with data from a Rwandan neonatal follow up clinic signifying that 46.5% of the 294 infants involved in the study were stunted, 19.9% were wasted and 44.2% were underweight.^5^Unfortunately, there is scarce information in Low and Middle -income countries (LMICs) pertaining to developmental delay during this same era of life. Prompt interventions like Kangaroo Mother Care (KMC), psychosocial support for parents, infant stimulation and individualized developmental care have been shown to positively impact weight gain, education and social-behavioral outcomes for these babies.^6-8^

Additionally, the first five years of life are extremely critical as they form a foundation for brain development and lifelong function.^9,10^ It is therefore important to intervene early with both educational and therapeutic services for those who are vulnerable.^10,11^ In order for this to happen, disabilities in development need to be identified early so that mitigating intervention can be started early leading to improved outcomes in the long-term.^12-14^Low birth weight (LBW), which includes prematurity, has been found to be a risk factor for acute and chronic malnutrition,^15^which significantly increases the risk of adverse neurodevelopmental outcomes such as gross motor, fine motor, language and social behavior in children born preterm.^6^

Infants born preterm are at risk of developing deficits in hearing and vision and the lower the gestation and the sicker the preterm neonate the higher the risk for these.^16-20^ Despite the fact that up to 6% of children born before the gestation of less than 26 – 27 weeks have severe to profound hearing loss, many lead normal or near normal lives after early intervention with cochlear implants.^21-23^ Visual problems of prematurity are many and on the whole those born with a birth weight of less than 1000 grams have a 3 times risk vis-a-vis the full-term of having a visual acuity of less than 6/60.^24,25^ The visual deficits of children born premature include cerebral visual impairment, ROP, refractive error, visual field defects, strabismus, colour vision deficits, decreased visual acuity (VA) and reduced contrast sensitivity (CS).^20,26^

Re-hospitalization after initial discharge from the newborn unit (NBU) is also high with rates ranging from 40.1% to 47.3% between discharge and the first year of life.^27,28^ Most of the re-admitted preterm infants are those delivered at a birth weight less than 1500 grams and before 33 weeks of gestation with chronic lung disease or complicated medical conditions and needs.^27,28^ The outcomes in development of preterm infants in the first year of life in this sub-Saharan region have not been extensively researched. However, a few studies provide relatable findings to the topic. For instance, Namazzi et al^6^conducted a study to establish the neuro-developmental results among premature infants and elicit any adjustable factors related to neuro-developmental disability (NDD). The study revealed a high incidence of NDD of 20.4% among preterm infants compared to 7.5% among the term babies of the same age. The most affected domain was fine motor (11.8%), followed by language (9.0%). At multivariate analysis, malnutrition and KMC at home after discharge were the key factors that were significantly associated with NDD among preterm babies.^6^

In Rwanda, a cross sectional study carried sought to explain the contrast in childhood development in children aged 24-36 months who were delivered premature and/ or low birth weight in comparison to their peers, and also evaluated factors related with suboptimal development. The children were 445 and 40 were preterm and/or LBW. 52.6% of the 445, with 27 (67.5%) of the preterm and/or LBW group, had developmental delay.^29^Were et al. conducted a study using the longitudinal cohort design to approximate the prevalence of neurological disabilities, as well as morbidity and mortality following discharge from Kenyatta National Hospital (KNH) at 2 years of age. Of the 120 infants evaluated, 14 (11.7%) had cerebral palsy, 11 (9.2%) were delayed on cognitive assessment while 32 (26.7%) were found to have functional disabilities. The factors associated with functional disability in the cohort included; neonatal illness, exclusive use of breast milk in the first month, neonatal weight gain less than 15 grams/kg/day, history of re-hospitalization and weight less than the third percentile at two years.^30^

Since neonatal care is being scaled up in Low and Middle -income countries (LMICs), it is important to understand in detail the state of the neonate post inpatient discharge and long term outcomes.^31,32^These include deaths, developmental impairments and malnutrition, which have been found to be more common in low birth weight neonates (LBW <2500 g).^15,33^ In Kenya, scale up of neonatal care is in part secondary to the efforts of NEST 360 (Newborn Essential Solutions and Technologies) alliance which is supporting Kenya, Malawi, Nigeria, and Tanzania to implement a health systems package for high-quality level-2 small and sick newborn care.^34^

Despite improvements in neonatal survival in Kenya, there is a paucity of data on neurodevelopmental, visual, and auditory outcomes in preterm infants and an eventual poor quality of care. Infants who are born small or sick are at risk of disability and poor development and so need additional attention to foster optimal development. Some of the outcomes experienced by the very low birth weight (VLBW) were studied 20 years ago and did not include vision and hearing.^30^ At the time of this study, new tools of neurodevelopment assessment like the ASQ and the Functional Near-Infrared Spectroscopy (fNIRS) had not been introduced into the country. fNIRS has become an increasingly feasible alternative and a potentially useful technique for studying functional activity of the infant brain as shown in several studies.^35-38^ There are therefore clinical gaps that need to be understood and incorporated into a comprehensive and follow-up programs.

This study aims to ascertain the prevalence of neurological disabilities such as motor impairments, cognitive and language delays, visual and hearing impairments, and behavioral and social-emotional challenges in preterm neonates 34 weeks’ gestation and less at the age of one year corrected for gestation following discharge from hospital NBUs and also to ascertain the factors associated with neurological disabilities at one year of age among these preterm neonates. It will also describe their growth patterns in the first year of life and explore their relationship to neurological outcomes. Their mortality will also be studied.

Our study is guided by the assumptions that preterm infants (<34 weeks) discharged from our local hospitals NBUs have a higher prevalence of neurodevelopmental, hearing, and visual impairments at one year of CA. Secondly, they have poor growth trajectories in the first year, which may be associated with worse neurodevelopmental outcomes. Finally, fNIRS can detect functional brain activity differences associated with developmental outcomes in preterm infants in this population.

As a result, this study will generate the first local evidence on the prevalence and possible determinants of neurodevelopmental disabilities in preterm infants, inform national guidelines on neonatal follow-up, and lead to the development of an evidence-based care bundle for comprehensive follow-up and early intervention. And the study will also contribute to the global literature on the feasibility of using the ASQ and fNIRS in LMIC contexts, as it is the first cohort study in Kenya that integrates neurological, growth, and sensory outcomes with this advanced imaging in preterm. The Kenyan neonates are currently managed as per the KNH Work Instructions for admission, transfer, discharge and follow up for the Newborn Unit Paediatrics Department (Appendix A).

## Methods

The study has been approved by KNH-UoN ethics research committee Reference Number P466/05/2023 and the Mbagathi County Referral Hospital. The parents or guardians of the participants will be informed about the purpose of the study and will receive oral and written information about it before participating. They will be given written informed consent to sign prior to recruitment and may withdraw from this study at any time without explanation. It will be stressed to them that if any disabilities and complications of prematurity are detected early, remedial or corrective services will be advised. Participant data will be handled confidentially and will be anonymized during publishing.

### Study Design

This is a hospital-based prospective cohort study of premature infants born at a gestation of 34 weeks or less discharged from the KNH and the Mbagathi County Referral Hospital NBUs until they attain one year corrected gestational age or death, whichever occurs first. They will be consecutively enrolled and followed prospectively from the point of discharge until 12 months of corrected age. Follow-up visits will occur at 40 weeks’ PMA or 2 weeks post discharge, 3, 6, 9, and 12 months, with phone calls to minimize loss to follow-up.

### Setting

The study will be conducted at the KNH and the Mbagathi County Referral Hospitals in Nairobi, Kenya. The KNH maternity unit handles over 9,000 deliveries a year. The KNH newborn unit has a Neonatal Intensive Care Unit (NICU) that caters for 2,400 neonates every year. The number of babies averages 70 to 100 daily, 85% of which are in born and 15% are referred from other health facilities or come from home. The Mbagathi County Referral Hospital maternity unit handles an average of 10,000 deliveries per year. The newborn unit has a NICU that caters for 2,200 neonates every year. The number of babies averages 60 to 80 daily, 70% of which are inborn and 30% are referred from other health facilities or come from home. These hospitals also have level-3 NICUs that manage small neonates (<1200 g/<30 weeks’ gestation) and offer mechanical ventilation and surgery. Currently the neonates are managed as per the KNH Work Instructions that include discharge and follow-up for the Newborn unit (Appendix A). Recruitment will be done sequentially on discharge after the mother or caretaker has given written informed consent.

## Enrollment and Eligibility Criteria

### Inclusion criteria

The Inclusion criteria will be premature neonates of gestation at birth of 34 weeks or less, whose mothers or guardians live in Nairobi city or its peri-urban areas (within a radius of 20 kms of the city center), and give written informed consent. The 20km radius limit is a reasonable distance that ensures proximity and better accessibility to the research sites, minimizes travel-related barriers that could increase the likelihood of dropouts, and captures a diverse population from the urban and peri-urban areas.

### Exclusion criteria

Infants with major congenital anomalies will be excluded as these deficits can affect neurological development. Congenital anomalies are anomalies that have significant medical, social or cosmetic implications on the affected individual and often require medical intervention; may be life threatening. For example, those of the Central nervous system (CNS) like hydro-cephalous and those of the Cardiovascular system (CVS).^39,40^

### Outcome variables

The proportion of survivors at one year of corrected gestational age determined by the ASQ and classified as any of the following; age appropriate development, borderline development or delayed development for each of the areas of communication, gross motor, fine motor, problem solving and personal-social.

The proportion of survivors at one year of corrected gestational age determined by fNIRS to have normal or reduced activity in the areas of the brain responsible for the following functions; motor (frontal lobe), vision (occipital lobe), speech (Broccas area), and cognition (prefrontal lobe). The proportion of survivors at one year of corrected gestational age who will be underweight, or have visual defects (strabismus, refractive errors, glaucoma and cataracts), or failed automated auditory brainstem response (AABR).

Gestational age is the duration of gestation measured from the first day of the last menstrual period expressed in completed weeks and days. The corrected gestational age is the age they would have been if born on their expected date of delivery. Chronological age is the age from the actual day the child was born.

### Primary Outcomes

The prevalence of neurological outcomes at one year of age in preterm neonates 34 week gestation and less discharged from the KNH and the Mbagathi County Referral Hospital NBUs defined by the ASQ and classified as any of the following; age appropriate development, borderline development or delayed development for each of the areas of communication, gross motor, fine motor, problem solving and personal-social.

The prevalence of visual defects (strabismus, refractive errors, glaucoma and cataracts), or failed automated auditory brainstem response (AABR) as determined by ophthalmological or visual assessment and Hearing screening.

The prevalence, as determined by fNIRS, of reduced or absent activity in the areas of the brain responsible for the following functions; motor (frontal lobe), vision (occipital lobe), speech (Broccas area), and cognition (prefrontal lobe).

### Secondary Outcomes

1. Growth of infants delivered prematurely at gestation of 34 weeks and less discharged from the KNH and the Mbagathi County Referral Hospital NBUs in the first year of life
2. Mortality in the first year of preterm infants delivered prematurely at gestation of 34 weeks and less discharged from the KNH and the Mbagathi County Referral Hospital NBUs.

### Sample Size

The sample size was determined by using Fisher’s formula for prevalence. An earlier study on neurological outcomes in preterm infants in KNH reported a prevalence of 30% neurological deficits (composite), mortality rate of 25.6% and loss to follow up rate of 5.6%.^38^ Therefore, a value of 30% was assumed and added to the original sample size yielding a sample size of 420.

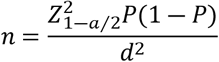

- Where, *n* is the sample size, 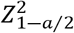 is the standard normal variate statistics corresponding to level confidence usually
- *Z* = 1.96 for 95% confidence interval, *P* is the expected proportion in population, and *d* is precision (margin of error)
- For the prevalence of 30%, and assuming 5% margin of error, the minimum sample size required will be:

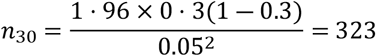

### Data Collection

Data collection is ongoing and commenced in April 2024. The process is expected to end in July 2026. Data pertaining to the mother is collected from the mother and child booklet, the mother’s files and from interviewing the mother. Data pertaining to the baby is collected from the baby’s referral and in-patient records. The anthropometric measures on discharge is performed by the clinical assistant. Subsequent data is obtained from the caretaker or mother’s history and from examination and evaluation at the clinic visits.

A pre-coded questionnaire is filled for each of these infants. Maternal data includes age, education level, occupation, menstrual history, ante-natal care, complications of pregnancy like ante-partum hemorrhage, pre-eclampsia, eclampsia, genito-urinary infections and hypertensive disease of pregnancy, place of birth and mode of delivery. Neonatal data includes birth weight, gestational age (as determined by the New Ballard’s score) and neonatal illnesses. Mobile phone numbers of mother/father/guardian and family is obtained. ROP screening is conducted before the infant is discharged or post discharge at 28 days of age for those who will have been discharged before 28 days of age.

### Follow-up Post-Discharge

Follow-up in both hospitals is at the neonatal outpatient clinic and this is depicted in the flow chart (Figure 1 -Flow chart of follow-up schedule). The first visit is at 2 weeks after discharge or 40 weeks post conception for those discharged before term corrected age. Subsequent visits are at every 3 months till the age of 1 year of CA. For those with complications visits are scheduled monthly as per the unit’s follow up criteria.

**Figure 1.**
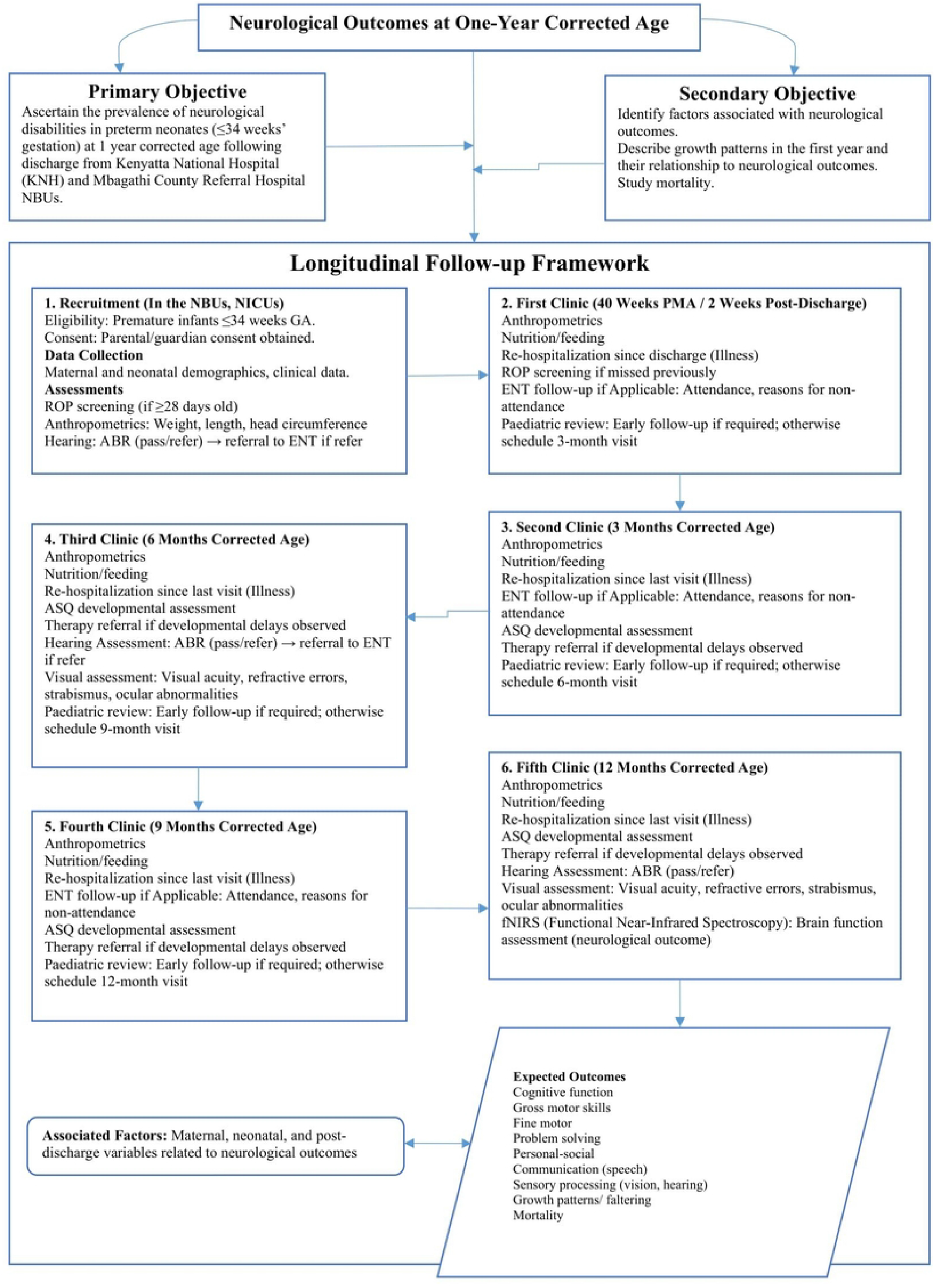

At every visit growth is monitored and the baby evaluated for weight, length and head circumference. Milestones are assessed every three months and any anomaly requiring occupational therapy attended to. Data pertaining deaths or re-hospitalization is recorded. At the age of 1 year the infants will undergo functional near infrared spectroscopy in order to determine and detect any reduced activity in the areas of the brain responsible for the following functions; motor (frontal lobe), vision (occipital lobe), speech (Broccas area), and cognition (prefrontal lobe). The infrared spectroscopy will be performed by a Paediatric neurodevelopmental specialist and will utilize the NIRSport 2™ machine. The machine is a wearable fNIRS that has eight dual wavelength Light Emitting Diode (LED) sources and eight detectors that offers a sampling rate of up to 7.81 Hz per channel. Data transmission is facilitated through Bluetooth^®^ and the device is powered by a rechargeable battery.

### Anthropometry Measurements

These is adopted from the INTERGROWTH-21^st^ International Fetal and Newborn Growth Standards for the 21st Century Anthropometry Handbook for the measurement of weight, height and head circumference of the preterm (Appendix B).

### Neurodevelopmental Assessment The ASQ

This is done using the ASQ as revised by the Human Development Center of the University of Oregon, which covers children aged from 1 to 66 months. It is filled by the parents or primary caregivers. The baby’s development for each of the areas of communication, gross motor, fine motor, problem solving and personal-social will be categorized as, on schedule (age appropriate development, needing learning activities), monitoring (borderline), needing further assessment with a professional (delayed development). This assessment will be at 3, 6, 9 and 12 months of age (Appendix C).

### Near Infrared Spectroscopy

In this study, for Functional Near Infra-red spectroscopy, the machine type NIRSport 2 by NIRx Medical Technologies will be used at the age of 1 year corrected for gestational age. This test will evaluate whether the functional activation in the targeted brain regions falls within the expected range for typical neurodevelopmental outcomes or a developing brain at this age, thus concluding whether the brain function is normal or reduced. The activity in the areas of the brain responsible for the following functions; motor (frontal lobe), vision (occipital lobe), speech (Broccas area), and cognition (prefrontal lobe) will be among our outcomes (Appendix C).

### Hearing Assessment

This is done using a handheld AABR (GSI Novus™). The GSI Novus is a sleek, handheld, comprehensive newborn hearing screening instrument. It features a touch screen display and intuitive software in a compact hardware design, and is classified as a class IIa device according to the European Union (EU) medical directive 93/42/EEC, and a class II device according to the United States Food and Drug Administration (US FDA). If the test is termed a fail, the infant is referred to an Audiologist for a diagnostic evaluation. If this test is normal a repeat will be performed at six months and at one year (Appendix C).

### Vision Assessment

All the eye examinations are conducted by a Paediatric Ophthalmologist stationed at the Kenyatta National Hospital and the Mbagathi County Referral Hospital. This is after the principal investigator has carried out the preliminary gross physical examination. The eye examination includes ROP screening, examination of the anterior segment, visual acuity determination, fundus examination and objective refraction (Appendix C).

### Reporting re-hospitalization

A readmission into a health facility for at least 24 hours will be labeled as a re-hospitalization. The parent/guardian will be requested to avail all documents with a record of the illness that will have occurred after the previous review. The details of the illness are recorded into the data form. Where these documents are not available the recall of the parent or guardian is the source of this data.

### Reporting Post-Discharge Mortality

The deaths that occur from the time of discharge from NBU until the end of the study will be recorded. At the initial contact the parent or guardian will be asked to notify the investigator of any infant deaths as soon as possible. This will be by mobile telephone or, physically at the next scheduled visit.

### Tracing Defaulters

For this purpose, the study has recruited a contact tracer who knows the Nairobi city geography well to visit the homes of defaulters not more than a month following a no show for a scheduled clinic appointment. Mobile phone contact is the other means by which missing subjects will be traced. All subjects will provide their mobile phone number and also one of a relative or close friend that can be contacted for information about their children whenever necessary.

## Data Analysis

### Descriptive Statistics

All maternal and neonatal baseline characteristics will be summarized using appropriate descriptive measures. Continuous variables will be reported as means with standard deviations (SD) or medians with interquartile ranges (IQR), depending on the normality of their distributions. Categorical variables will be summarized using frequencies and percentages.

The percentage of study participants who will not have attained the 10^th^ percentile of the WHO anthropometric measures by term at 12 months corrected age will be reported among those surviving to 1 year. Similarly, the prevalence of neurological deficits, including visual (e.g., strabismus, refractive errors, glaucoma, cataracts), hearing, and developmental delays as determined by ASQ will be presented as proportions of one-year survivors. In addition, the prevalence of altered functional activity as determined by fNIRS measurements targeting brain regions associated with motor (frontal lobe), vision (occipital lobe), speech (Broca’s area), and cognition (prefrontal cortex) will be summarized as percentages.

### Comparative Analysis

Growth parameters measured at multiple time points (3, 6, 9, and 12 months) will be analyzed and compared across subgroups defined by sex, breastfeeding status, intrauterine growth, infant illnesses, and maternal illnesses. For continuous variables (e.g., growth measurements), appropriate statistical tests will be applied to compare medians or means, depending on data distribution. Outcomes of interest in these analyses include postnatal growth, as well as visual, hearing, and neurological measures. For categorical variables, cross-tabulations and Chi-square tests will be used to calculate relative risks (RR), with corresponding p-values and 95% confidence intervals, to assess associations between exposure factors and the outcomes.

### Modelling approaches

To assess growth delays and determine factors affecting this outcome, growth measurements and neurological deficits will be evaluated at multiple time points (3, 6, 9 and 12 months). Mixed-effects models will be used as the primary analytic approach. Mixed-effects models are particularly appropriate for this objective because they accommodate the correlation of repeated measurements within the same infant and allow for individual-level random effects, enabling the modelling of each infant’s trajectory over time. In contrast, a single-time-point logistic regression model would treat each measurement as independent, ignoring within-subject correlation and thus potentially yielding biased estimates and incorrect standard errors.

Generalized Estimating Equations (GEE) will be used as part of sensitivity analysis to confirm the robustness of the results. GEE provides population-averaged estimates and is less sensitive to the specification of the random-effects structure, offering a complementary perspective.

For the mortality outcome, the time-to-event (survival) modelling approach will be applied. Kaplan-Meier curves will be constructed to estimate survival probabilities over the first year of life, and log-rank tests will compare survival functions across subgroups. To determine how covariates affect the time to experience event of interest (death), Cox-proportional hazards regression models will be fitted to identify predictors of mortality, providing hazard ratios and 95% confidence intervals.

### Handling missing data

All efforts will be made to minimize missing data. Where missing data are present, multiple imputations with chained equations (MICE) will be considered under the assumption of data missing at random. Sensitivity analyses will be conducted to assess the robustness of findings.

All analyses will be done using R programming language.

### Study Timeline

Recruitment and follow up are ongoing and occurring concurrently in this study. Recruitment began on 3^rd^ April 2024. The processes will end on 3^rd^ September 2026. Data collection will end shortly afterwards 3^rd^ October 2026, with cleanup starting after that. After the data is cleaned for an estimated period of one month, it will be prepared for analysis, with results expected in December 2026.

## Discussion

The preterm infant is at risk of adverse outcomes post-inpatient discharge. These include deaths, developmental impairments and malnutrition, which have been found to be more common in low birth weight neonates (LBW <2500 g).^15,33^ The developmental impairments include cerebral palsy, ROP, hearing and seeing deficits among others. It is therefore important that these infants be closely monitored and screened for early identification and mitigation.

These post-inpatient/long-term outcomes in the first year of life have not been extensively researched in this sub-Saharan region. Some of these outcomes were studied in Kenya in a limited fashion more than 20 years ago. During this period, neonatal care has been scaled up with the introduction of Continuous Positive Airway Pressure (CPAP), surfactant and mechanical ventilation that has led to the survival of preterm infants. The ASQ and the fNIRS methods of assessing development have only recently been introduced into Kenya. It is projected that this study will assess more extensively the current post-inpatient discharge and long-term outcomes of our preterm and that the results from this study will lead to a comprehensive and affordable protocol for the follow-up of these vulnerable infants that can be utilized even in the lower level of care centers. This also aligns with United Nations Sustainable Development Goal (SDG) 3.2, which targets ending preventable deaths of newborns and children under five and ensuring that every child has a chance to thrive.

The strengths of this study is that it is the first in Kenya in preterm infants that incorporates visual, hearing and neurodevelopmental follow-up, the first to utilize the ASQ mode of developmental assessment, and to assess neurodevelopment utilizing functional near infrared spectroscopy. The main limitation is of dropouts during follow up, which has been taken care of in the sample calculation and is mitigated through physical tracing and cellular contact. The results of the study will be published in conference abstracts and presentations and peer-reviewed articles in scientific journals.

## Data Availability

No datasets were generated or analysed during the current study. All relevant data from this study will be made available upon study completion.

## Data Availability

No data are associated with this article.

## Author Contributions

1. Florence Murila (Corresponding author) – was involved in conceptualization, data curation, investigation, design of methodology, project administration, provision of resources and writing of the original draft.
2. Fred Were – was involved in conceptualization, supervision and review and editing.
3. Jalemba Aluvaala - was involved in conceptualization, design of methodology, supervision and review and editing.
4. Moses Obimbo - was involved in conceptualization, supervision and review and editing.

